# Predictors of tobacco use among the youth in an urban slum in Kampala, Uganda: A cross-sectional study

**DOI:** 10.1101/2025.03.06.25323538

**Authors:** Joyce Nakitende, Denis Omara, Dennis Kalibbala, Geofrey Musinguzi, Bontle Mbongwe

## Abstract

**Background and justification:** Tobacco use remains a significant public health concern worldwide and is the leading risk factor for developing non-communicable diseases. In Uganda, the youth use tobacco at three times the rate of adults, with those residing in slum areas exhibiting even higher prevalence levels. Since 2015, strict laws regulating public tobacco use have been implemented in Uganda, however, these measures have not led to a significant decline in tobacco consumption among the youth in slums.

**Objective:** To assess the predictors of persistent tobacco use and associated factors among youth living in the slum areas of Kampala, Uganda.

**Methods:** This cross-sectional study utilized mixed methods of data collection. Conducted in Bwaise slum of Kampala, the study recruited 422 youths aged 18-30 years. Households were sampled systematically, and quantitative data were analyzed using STATA version 14. Logistic regression was used, Odds ratios were used to measure the associations. Factors were considered significant at p<0.05

**Results:** The prevalence of current tobacco use was 52.6% while the prevalence of ever use was 71.6%. The majority (87.4 %) of the participants knew the health effects of tobacco use. Males were more than twice as likely to smoke compared to females (aOR: 2.72, 95% CI: 1.66-4.44). Participants aged 21 years and above were over twice as likely to smoke compared to those 20 years and below (aOR: 2.54, 95% CI: 1.45-4.45). Additionally, participants who were unaware that smoking causes serious illness were four times more likely to smoke compared to those who were knowlegeable (aOR: 4.49, 95% CI: 1.15-17.56).

**Conclusion:** More than half of the youth use tobacco despite awareness of its health effects. Male gender aged 21-30 years and lack of knowledge regarding the serious illness caused by smoking were strongly associated with tobacco use. This calls for development and implementation of targeted educational initiatives that address the unique needs and behaviors of males aged 21 to 30 years.

## Introduction

Globally, tobacco-related deaths have been estimated to exceed 8 million annually (1) According to the World Health Organization (WHO) and the Centers for Disease Control and Prevention (CDC), this figure is projected to rise by the year 2030 (2), with Sub-Saharan Africa expected to be disproportionately affected (3). In Uganda, approximately 10% of the population uses tobacco daily, with usage rates among the youth and young adults being significantly higher, sometimes doubling or tripling (4, 5).

The most commonly used forms of tobacco include water pipe tobacco, various smokeless tobacco products, cigars, cigarillos, roll-your-own tobacco, pipe tobacco, bidis, kreteks, kuber, shisha among others (6). Unlike conventional smoking, users of smokeless tobacco often consume these without detection by others in public spaces (6) making comprehensive control of their use more challenging.

Tobacco use is recognized as the leading and most significant risk factor associated with non-communicable diseases (NCDs) and other adverse health outcomes such as hypertension, oral cancers, lung cancers, chronic obstructive pulmonary disease (COPD), adverse reproductive health outcomes and sudden infant death syndrome (SIDS) (7). Most smokers initiate their habit during adolescence and evidence suggests that as they transition into adulthood, overcoming addiction increasingly become difficult (8). In Uganda, 10.5% of youth aged 13-15 used any tobacco product in 2018 (9). Furthermore, a recent study reported a high prevalence of 36% among the individuals aged 18-30 years who smoke Shisha, one of the most common and emerging tobacco products (4).

To address the global tobacco epidemic, the World Health Organization (WHO) has identified six evidence-based measures collectively referred to as “MPOWER” outlined in the Framework Convention on Tobacco Control (FCTC) (1). These measures include monitoring tobacco use and prevention policies, protect people from tobacco smoke, offering assistance to quit tobacco use, warn the public about the dangers of tobacco, enforcing bans on tobacco advertising, promotion and sponsorship as well as raising taxes on tobacco.

In 2015, the Ugandan Ministry of Health enacted legislation that aligns with the FCTC and incorporates the essential components of the MPOWER framework. The implementation of this law is currently on ongoing (10–12). However, several nationwide studies focusing youth have reported no significant decline in tobacco use (9) and in some instances, an increase in prevalence compared to previous years (4, 5).

Youth living in slums are particularly vulnerable to tobacco use largely due to the socio-economic challenges they face (13). In Uganda, over 70% of the population consists of the youth with over 60% of the population in Kampala residing in slum conditions(14). The continued rise in tobacco use among this demographic, despite the presence of stringent laws warrants closer examination.

This study aims to assess the predictors of persistent tobacco use and the associated factors among youth living in a slum in Kampala, Uganda. Understanding these dynamics is crucial for developing targeted interventions that can effectively reduce tobacco consumption in this high-risk population.

## Methods

### Study site

This study was conducted within Bwaise, the largest slum settlement in Kampala, the capital city of Uganda. Kampala is divided into 5 divisions namely: Central, Makindye, Kawempe, Nakawa and Rubaga divisions. Approximately 60% of Kampala’s population resides in slum areas with a total of 57 slums distributed across these divisions (14). Bwaise alone contains over 20,000 households and is divided into 3 large parishes each averaging around 7000 households. Bwaise is characterized by extreme poverty, high unemployment rates, elevated crime levels, inadequate sanitation and hygiene, as well as issues related to illicit drug use and prostitution among others (15).

### Study population

The study was conducted among the youth aged 18-30 who are residents of Bwaise, in Kampala city. While the United Nations (UN) defines a youth as a person between 15-24 years (16) this study adhered to the definition provided by the constitution of Uganda, which identifies youth as those aged 18-30 years (17).

### Study design

This was a prospective cross-sectional study in which quantitative data were collected.

### Sample size estimation

Kish Leslie’s (1965) formula for proportions (18) was utilized in this study to determine the required sample size. The formula is expressed as:

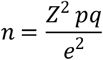

Where n represents the required sample size, Z=1.96 is the standard normal value corresponding to a 95% confidence level, p is the estimated prevalence of tobacco use among the youth residing in the slums of Kampala (set at 50%), q is defined as (1-p), and e is the desired level of precision (0.05).

Substituting the values into the formula above; 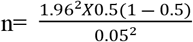 n = 348 participants

Considering an anticipated non-response rate of 10%, the adjusted sample size was calculated as: n = 384/(1-0.10) n = 422 Participants. Thus, a total of 422 participants were targeted for the study to account for potential nonresponse.

### Sampling technique

Bwaise was selected purposively as it is the largest slum in Kampala district. Within Bwaise, one zone was randomly chosen, and the households within that zone were systematically sampled to recruit the eligible participants for the study.

### Sampling households

Recruitment and data collection started on 30^th^ March 2021 and ended on 21^st^ April 2021. Bwaise slum is divided into 3 parishes, each containing approximately 7000 households. One of these parishes was selected randomly for the study. Within the chosen parish, the first household was also selected randomly. Initially, a systematic sampling approach was employed with the intention of sampling every 16^th^ household to ensure a representative sample. The first approximately 20 households were sampled systematically at this interval. However, given the informal nature of the setting, subsequent households were sampled conveniently until the target sample of 422 was achieved. This approach allowed for a practical adaptation to the challenges presented by the slum environment while still aiming to reach the desired sample size.

### Study variables

#### The dependent variable

Tobacco use served as the dependent variable in this study and was measured using standardized questions adapted from the Global Adult Tobacco Survey (GATS) and the WHO STEP-wise instrument (19, 20). According to the GATS, tobacco use is classified into categories of current tobacco users, which includes subcategories such as daily smokers, occasional smokers and former daily or occasional smokers. Non-tobacco users are further categorized into former daily smokers, never daily smokers and former occasional smokers. In this study, we specifically assessed both current and ever tobacco use among participants.

#### The Independent variables

Independent variables in this study included participants’ social demographics, and their knowledge of the health effects of tobacco use amongst the youth. Social demographics encompassed factors such as age, gender, education level, income, and occupation. Knowledge of the health effects of tobacco use was assessed through questions designed to evaluate participants’ awareness and understanding of the risks associated with tobacco consumption.

### Data analysis

All data were entered into EpiData version 4.1 and subsequently exported to STATA version 14 for analysis. Tobacco use patterns and prevalence were assessed through frequency counts. The results for this objective were presented using measures of central tendency and dispersion including mean, mode and standard deviation.

To evaluate significant associations between knowledge of tobacco use and social demographics, logistic regression analyses were conducted at both bivariate and multivariate levels. Factors with a p-value of less than 0.05 were considered statistically significant.

### Ethical considerations

Approval to conduct the study was obtained from the Higher Degrees Research Ethics Committee at Makerere University School of Public Health (approval is attached as additional information). Further permission was obtained from the local community leaders.

Written informed consent was obtained from every eligible participant.

## Results

### Characteristics of the study participants

A total of 422 youths from Bwaise participated in the study with 248 (58.8%) being male. The mean age of participants was 23 years (±3.64 Standard Deviation [SD]) with a majority (70.8%) aged 21-30 years. On average participants had 9.6 (SD±3.27) of schooling while there were 3(±2) people on average older than 18 years living in the house (Table 1). Approximately 80% of participants were employed. With over 42% earning less than 250,000/month.

**Table 1.** Characteristics of the study participants.

|  | Number of respondents (n) | Proportion (%) |
| --- | --- | --- |
| **Gender** |  |  |
| Male | 248 | 58.8 |
| Female | 174 | 41.2 |
| **Age, Mean(SD)** | **23($\pm 3.64$)** |  |
| 18-20years | 123 | 29.2 |
| 21-30years | 299 | 70.8 |
| **Marital status** |  |  |
| Not married | 314 | 74.4 |
| Married | 108 | 25.6 |
| **Region of origin** |  |  |
| Central | 305 | 72.3 |
| Eastern | 41 | 9.7 |
| Western | 58 | 13.7 |
| Others | 18 | 4.3 |
| **Education level** |  |  |
| None/primary | 137 | 32.5 |
| Secondary/University | 285 | 67.5 |
| **Religion** |  |  |
| Christian | 226 | 53.6 |
| Non-Christian | 196 | 46.4 |
| **Employment status** |  |  |
| Employed | 336 | 79.8 |
| Unemployed | 45 | 10.7 |
| House wife | 18 | 4.3 |
| Students | 22 | 5.2 |
| **Income/month (UGX)** |  |  |
| less than 250,000/ | 177 | 41.9 |
| 250,000-500,000/ | 90 | 21.4 |
| 500,000-750,000/ | 25 | 5.9 |
| 750,000-1,000,000/ | 14 | 3.3 |
| Above 1,000,000/ | 116 | 27.5 |

**Table 2.** Tobacco use characteristics of the study participants.

|  | Frequency<br>(n) | Percentage<br>(%) |
| --- | --- | --- |
| **Currently smoke tobacco products, such as cigarettes, cigars or pipes** |  |  |
| Yes | 222 | 52.6 |
| No | 200 | 47.4 |
| **Currently smoke tobacco products daily (n=222)** |  |  |
| Yes | 160 | 72.1 |
| No | 62 | 27.9 |
| **Type of cigarette (n=222)** |  |  |
| Manufactured cigarettes | 95 | 42.8 |
| Hand-rolled cigarettes | 18 | 8.1 |
| Pipes full of tobacco | 48 | 21.6 |
| Cigars, cheroots, cigarillos | 4 | 1.8 |
| Shisha | 122 | 55.0 |
| “Kibanga” | 115 | 51.8 |
| **During the past 12 months, have you tried to stop smoking (n=222)** |  |  |
| Yes | 128 | 57.7 |
| No | 94 | 42.3 |
| **Advised to quit smoking tobacco by a health worker in the past 12 months (n=222)** |  |  |
| Yes | 36 | 16.2 |
| No | 66 | 29.7 |
| No visit during the past months | 120 | 54.1 |
| **In the past, did you ever smoke any tobacco products (show card)** |  |  |
| Yes | 80 | 40.0 |
| No | 120 | 60.0 |
| **Currently use any smokeless tobacco such as snuff, chewing tobacco, betel** |  |  |
| Yes | 69 | 16.4 |
| No | 353 | 83.6 |
| **Currently use smokeless tobacco products daily** |  |  |
| Yes | 44 | 63.8 |
| No | 25 | 36.2 |
| **Ever used smokeless tobacco in the past** |  |  |
| Yes | 126 | 29.9 |
| No | 296 | 70.1 |
| **Ever use smokeless tobacco daily in the past** |  |  |
| Yes | 53 | 42.1 |
| No | 73 | 57.9 |
| **During the past 30 days, did someone smoke in your home?** |  |  |
| Yes | 194 | 46.0 |
| No | 228 | 54.0 |
| **During the past 30 days, someone smoked in closed areas at workplace** |  |  |
| Yes | 322 | 76.3 |
| No | 62 | 14.7 |
| Don't work in a closed area | 38 | 9.0 |

**Table 3.** Knowledge about the health effects of tobacco use among the study participants.

|  | Number of respondents (n) | Proportion (%) |
| --- | --- | --- |
| **Knows or believes that smoking tobacco causes serious illness** |  |  |
| Yes | 369 | 87.4 |
| No | 30 | 7.1 |
| Don't know | 23 | 5.5 |
| **Knows or believes that smoking tobacco causes Stroke (blood clots in the brain that may cause paralysis)** |  |  |
| Yes | 189 | 44.8 |
| No | 85 | 20.1 |
| Don't know | 148 | 35.1 |
| **Knows or believes that smoking tobacco causes Heart attack** |  |  |
| Yes | 278 | 65.9 |
| No | 64 | 15.1 |
| Don't know | 80 | 19.0 |
| **Knows or believes that smoking tobacco cause Lung cancer** |  |  |
| Yes | 383 | 90.8 |
| No | 14 | 3.3 |
| Don't know | 25 | 5.9 |
| **Knows or believes that using \*smokeless tobacco\* causes serious illness** |  |  |
| Yes | 305 | 72.3 |
| No | 41 | 9.7 |
| Don't know | 76 | 18.0 |

**Table 4.** Current smoker and ever-smoked associations with knowledge among the youth in Bwaise.

|  | Current Smoker |  |  | Ever Smoked |  |  |
| --- | --- | --- | --- | --- | --- | --- |
|  | Males<br>(n= 158)<br>n (%) | Females<br>(n= 64)<br>n (%) | p-value | Males<br>(n=208)<br>n (%) | Females<br>(n=94)<br>n (%) | p-value |
| **Knows or believes that smoking tobacco cause serious illness** |  |  | 0.112 |  |  | 0.105 |
| Yes | 126(79.7) | 53(82.8) |  | 174(83.7) | 82(87.2) |  |
| No | 23(14.6) | 4(6.3) |  | 23(11.1) | 4(4.3) |  |
| Don't know | 9(5.7) | 7(10.9) |  | 11(5.3) | 8(8.5) |  |
| **Knows or believes that smoking tobacco causes Stroke (blood clots in the brain that may cause paralysis)** |  |  | 0.642 |  |  | 0.563 |
| Yes | 64(40.5) | 24(37.5) |  | 89(42.8) | 41(43.6) |  |
| No | 45(28.5) | 16(25.0) |  | 53(25.5) | 19(20.2) |  |
| Don't know | 49(31.0) | 24(37.5) |  | 66(31.7) | 34(36.2) |  |
| **Knows or believes that smoking tobacco causes Heart attack** |  |  | 0.791 |  |  | 0.548 |
| Yes | 93(58.9) | 39(60.9) |  | 134(64.4) | 64(68.1) |  |
| No | 36(22.8) | 12(18.8) |  | 37(17.8) | 12(12.8) |  |
| Don't know | 29(18.3) | 13(20.3) |  | 37(17.8) | 18(19.1) |  |
| **Knows or believes that smoking tobacco cause Lung cancer** |  |  | 0.501 |  |  | 0.276 |
| Yes | 137(86.7) | 53(82.8) |  | 186(89.4) | 81(86.2) |  |
| No | 9(5.7) | 3(4.7) |  | 10(4.8) | 3(3.2) |  |
| Don't know | 12(7.6) | 8(12.5) |  | 12(5.8) | 10(10.6) |  |
| **Knows or believes that using smokeless tobacco causes serious illness** |  |  | **0.017\*** |  |  | 0.095 |
| Yes | 112(70.9) | 35(54.7) |  | 147(70.7) | 58(61.7) |  |
| No | 21(13.3) | 8(12.5) |  | 26(12.5) | 10(10.6) |  |
| Don't know | 25(15.8) | 21(32.8) |  | 35(16.8) | 26(27.7) |  |

**Table 5.** Univariate and multivariate analysis of factors associated with tobacco use.

|  | Univariate analyses | Multivariate analyses |  |  |
| --- | --- | --- | --- | --- |
|  | All | All | Male | Female |
|  | OR(95% CI) | aOR(95% CI) | aOR(95% CI) | aOR(95% CI) |
| **Gender** |  |  |  |  |
| Female | 1 | 1 |  |  |
| Male | **3.02(2.02-4.51)** | **2.72(1.66-4.44)** | - | - |
| **Age** |  |  |  |  |
| 18-20years | 1 | 1 | 1 | 1 |
| 21-30years | **2.34(1.36-4.04)** | **2.54(1.45-4.45)** | **2.12(1.07-4.23)** | **4.51(1.40-14.55)** |
| **Marital status** |  |  |  |  |
| Not married | 1 | 1 | 1 | 1 |
| Married | 1.01(0.65-1.56) | 1.03(0.60-1.75) | 1.81(0.87-3.78) | 1.51(0.20-1.32) |
| **Region of origin** |  |  |  |  |
| Central | 1 | 1 | 1 | 1 |
| Outside Central | **0.58(0.38-0.88)** | 0.84(0.60-1.18) | 0.69(0.33-1.42) | 0.81(0.35-1.89) |
| **Education level** |  |  |  |  |
| None/primary | 1 | 1 | 1 | 1 |
| Secondary/University | **0.54(0.35-0.82)** | 0.64(0.39-1.04) | 0.71(0.37-1.37) | 0.42(0.16-1.04) |
| **Religion** |  |  |  |  |
| Christian | 1 | 1 | 1 | 1 |
| Non-Christian | 1.30(0.88-1.91) | 1.42(0.89-2.27) | 1.62(0.90-2.91) | 1.46(0.59-3.59) |
| **Employment status** |  |  |  |  |
| Employed | 1 | 1 | 1 | 1 |
| Unemployed | 0.60(0.32-1.12) | 1.11(0.51-2.39) | 1.14(0.34-3.79) | 1.13(0.35-3.64) |
| House wife | 0.34(0.13-1.02) | 0.42(0.11-01.51) | - | 0.45(0.09-2.38) |
| Students | **0.12(0.03-0.41)** | 0.39(0.10-1.62) | 0.91(0.12-7.05) | 0.21(0.02-2.73) |
| **Income** |  |  |  |  |
| less than 250,000/= | 1 | 1 | 1 | 1 |
| 250,000-500,000/= | 0.68(0.41-1.13) | 0.55(0.30-1.02) | 0.65(0.30-1.39) | 0.28(0.08-1.04) |
| 500,000-750,000/= | 0.49(0.21-1.14) | 0.41(0.21-1.10) | 0.44(0.15-1.30) | 0.21(0.02-2.61) |
| 750,000-1,000,000/= | 0.34(0.11-1.08) | 0.28(0.11-1.08) | 0.27(0.07-1.08) | - |
| Above 1,000,000/= | **0.47(0.29-0.76)** | 0.71(0.40-1.27) | 0.74(0.32-1.68) | 0.61(0.24-1.54) |
| *OR = Odds Ratio*<br>*aOR = adjusted Odds Ratio*<br>*CI = Confidence Interval* |  |  |  |  |

### Tobacco use characteristics of the study participants

The majority of the youth, 302 (71.6%) had ever smoked tobacco products. Current smokers of tobacco products such as cigarettes, cigars or pipes were 222 (52.6%). The mean age at start of smoking was 17 years (SD= ± 3.91). A big proportion (57.7%) of the current smokers had tried to stop smoking in the past 12 months.

### Knowledge about the health effects of tobacco use among the study participants

Overall, the majority of participants knew the effects of tobacco use on health. More than 3/4 knew that tobacco causes serious illnesses, lung cancer and that smokeless tobacco causes serious illnesses. While less than 1/2 knew that smoking tobacco causes stroke.

### Current smoker and ever-smoked associations with knowledge variables

Knowing or believing that using smokeless tobacco causes serious illness was statistically significantly associated with current tobacco use among both males and females.

### Factors associated with tobacco use

A multivariate analysis showed that males were 2.7 times more likely to smoke than females. (aOR: 2.72, 95% CI: 1.66-4.44). Participants aged 21-30 years were 2.5 times likely to smoke than those aged 18-20 years (aOR: 2.54, 95% CI: 1.45-4.45).

### Table 6: Additional multivariate analysis of factors associated with tobacco use

Participants who didn’t know or believe that smoking tobacco cause serious illness were 4.49 times likely to smoke (aOR: 4.49, 95% CI: 1.15-17.56) and this was significantly associated with tobacco use. Females who didn’t know that smokeless tobacco causes serious illness were 8.87 times likely to smoke compared to their male counterparts.

**Table 6.** Additional multivariate analysis of factors associated with tobacco use.

|  | Univariate analyses | Multivariate analyses |  |  |
| --- | --- | --- | --- | --- |
|  | All | All | Male | Female |
|  | OR(95% CI) | aOR(95% CI) | aOR(95% CI) | aOR(95% CI) |
| **Knows or believes that smoking tobacco causes serious illness** |  |  |  |  |
| Yes | **1** | **1** | **1** | **1** |
| No | **9.55(2.84-32.04)** | **4.49(1.15-17.56)** | **7.04(1.27-39.16)** | 7.38(0.45-120.81) |
| Don't know | **2.43(0.97-6.04)** | 1.15(0.40-3.35) | 1.14(0.28-4.53) | 0.67(0.10-4.49) |
| **Knows or believes that smoking tobacco causes Stroke (blood clots in the brain that may cause paralysis)** |  |  |  |  |
| Yes | **1** | **1** | **1** | **1** |
| No | **2.92(1.68-5.07)** | 1.63(0.84-3.14) | 1.30(0.58-2.89) | 2.17(0.59-7.94) |
| Don't know | **1.12(0.72-1.72)** | 0.87(0.51-1.46) | 0.88(0.44-1.80) | 0.78(0.31-1.98) |
| **Knows or believes that smoking tobacco causes Heart attack** |  |  |  |  |
| Yes | **1** | **1** | **1** | **1** |
| No | **3.32(1.80-6.12)** | 1.91(0.91-4.03) | 1.40(0.58-3.35) | 3.10 (0.63-15.20) |
| Don't know | 1.22(0.74-2.01) | 0.84(0.47-1.51) | 0.76(0.35-1.65) | 1.33(0.43-4.12) |
| **Knows or believes that smoking tobacco causes Lung cancer** |  |  |  |  |
| Yes | **1** | **1** | **1** | **1** |
| No | **6.09(1.34-27.59)** | 1.19(0.20-7.18) | 0.60(0.09-4.09) | - |
| Don't know | **4.06(1.49-11.04)** | **3.81(1.18-12.26)** | 4.21(0.77-23.07) | 3.46(0.54-21.95) |
| **Knows or believes that using smokeless tobacco causes serious illness** |  |  |  |  |
| Yes | **1** | **1** | **1** | **1** |
| No | **2.60(1.28-5.28)** | 2.13(0.90-5.01) | 1.12(0.41-3.08) | **8.87(1.71-45.95)** |
| Don't know | 1.65(0.98-2.75) | 1.82(0.96-3.45) | 0.79(0.33-1.86) | **5.86(1.90-18.07)** |
| *OR = Odds Ratio*<br>*aOR = adjusted Odds Ratio*<br>*CI = Confidence Interval* |  |  |  |  |

## Discussion

In this study, more than 5 in 10 youths who dwell in this slum used tobacco. Approximately similar results were obtained in Uganda in a survey of patients living with HIV (21), among males who work in a police barracks (22), and among pregnant women in Maracha district (23). Similarly in Palestine, the reported prevalence of tobacco use was 47.7% (24), and 42.3% in a study among young men in Bangladesh (25). On the contrary, this is slightly higher than the prevalence reported by the recent estimates for Uganda in the GYTS survey (5). Similarly, the GATS estimates for Uganda reported 1 in 10 prevalence of smoking in the general population (4, 26). This can be attributed to many factors: For the estimates from GYTS (Uganda), the definition of youths was different from that used in our study (Ugandan constitution definition of a youth (18–30) were included in our study. For the estimates from the GATS, that survey was not specific to slum areas but rather to the general Ugandan population. The high level of prevalence in our study can thus be attributed to the nature of the study site, a slum setting characterized by high crime rate, poverty in extremes, and rampant use of drugs and

### The most common type of tobacco used

Unlike in most studies done worldwide (24, 27), the most common form of tobacco used was termed as “*Kibanga*”, a local name used to mean a mixture of tobacco and marijuana, hand rolled to form a stick of mixed substance. From our qualitative interviews, yet to be published, some participants mentioned that this mixture of 2 substances (marijuana and tobacco) was safe, neutral and had no effects on human health whatsoever. An informed participant said that they were aware that tobacco has ill health effects, however, marijuana having documented health benefits neutralizes those ill effects caused by tobacco, hence the mixture can be consumed in huge amounts, however, there is scanty literature on this.

We also found out that several participants who consumed tobacco at most times narrated to have firstly had chewed mira-a form of leaf abused in Uganda. infact some reported that each time they chewed mira, it automatically led them to smoke tobacco,” mira asks you for smoke” which is then used in form of tobacco. Similar findings were documented in other studies (28–30).

Males in this study were 3 times more likely to smoke compared to females. Evidence globally suggests that males have higher smoking levels compared to women, mostly linked to peer pressure and social reasons (31–33). Similar evidence was documented in the recent GATS survey in Uganda (26). On the other hand, there is very scanty literature suggesting otherwise.

Generally, the majority of the participants in this study had good knowledge about the ill effects of tobacco, much as they were smokers. These findings are consistent with those from Bangladesh (34) and New Delhi (35). Persistent tobacco use even when one is aware of its ill effects may be explained by several cessation barriers such as lack of social support and continued peer pressure. Of importance was the finding that participants who didn’t know or believe that smoking causes serious illness were 4.49 times more likely to smoke than their counterparts. This can be linked to the fact that the more one perceives oneself to be at less risk, the more they continue to smoke. The WHO advises that there are still more opportunities to cover smoking knowledge gaps (36).

## Limitations

This was a cross-sectional study, it couldn’t infer causality. The data was based on self-reports, which is subject to response bias

## Conclusion

More than half of the youth use tobacco despite knowing the health effects of tobacco use. Being male aged 21-30 years and unaware of the serious illness caused by smoking tobacco was strongly associated with tobacco use.

## Data Availability

All relevant data are within the manuscript and its Supporting Information files.

## Acknowledgements

We acknowledge the study participants who provided the data and Bwaise local leaders.

## Author contributions

Conceptualization: JN, GM; Methodology: JN, GM, BM; Investigation: JN, GM; Formal analysis: JN, DK, BM; Funding acquisition: JN; Writing, Review and editing: JN, DO, GM, BM

